# IL-17/IL-10 ratio and Viral Load changes in HIV-1 Patients on First-Line Antiretroviral Therapy at Moi Teaching And Referral Hospital, Kenya

**DOI:** 10.64898/2025.12.03.25339332

**Authors:** Rose Undisa, Isacc Ndede, Lameck Diero

## Abstract

**Background:** Persistent low-level viremia (pLLV) in HIV-1 patients on combination antiretroviral therapy (cART) poses a clinical challenge, potentially driving immune activation and increasing the risk of virologic rebound. While the IL-17/IL-10 ratio plays a key role in immune regulation, its significance in pLLV remains under-explored, particularly in sub-Saharan Africa. This study assessed the variations in viral load (VL), with IL-17/IL-10 ratio in HIV-1 patients on first-line cART at Moi Teaching And Referral Hospital, Kenya.

**Objective:** To determine the viral load and evaluate the IL-17/IL-10 ratio in HIV-1 patients on first-line cART with persistent low-level viremia compared to virally suppressed patients at Moi Teaching And Referral Hospital, Kenya.

**Methodology:** This cross-sectional study recruited 82 HIV-1 patients on first-line cART, 41 pLLV (VL 50–500 copies/ml), and 41 virally suppressed participants (VL <50 copies/ml), age and gender matched 1:1. Patients were purposively sampled from the Moi Teaching And Referral Hospital, Medical Records System (MRS) from January 2021 to December 2022. Blood samples were collected in EDTA and analysed for IL-17 and IL-10 via ELISA (Zeptometrix, USA). Viral loads were determined using Abbott Real-Time PCR (version 4.0, USA). The IL-17/IL-10 ratios were calculated to evaluate immune balance. Data was analyzed using STATA version 15. Mann-Whitney U-test was used to compare ratios between the groups; p ≤ 0.05 was considered statistically significant.

**Results:** The median (IQR) VL in the studied HIV-1 patients with pLLV was 407.5 (314.5-445.0) copies/ml. HIV-1-suppressed patients had a slightly higher mean IL-17/IL-10 ratio of 0.556 vs 0.552 in pLLV, P=0.433, and was positively correlated (rho= 0.453, p=0.003) to reduced viral load and hence viral suppression. The odds (95% CI) of being suppressed were decreased by IL-17, 0.938(0.691-1.273), but increased by IL-10, 1.106(0.675-1.811), though not statistically significant.

**Conclusion:** The study found no statistically significant difference in IL-17/IL-10 ratios between pLLV and virally suppressed HIV-1 patients on cART. While these findings suggest a possible involvement of IL-17/IL-10 immune balance in viral control, they remain exploratory. Further research with larger cohorts is needed to clarify these relationships and to determine whether cytokine biomarkers could complement existing tools for monitoring treatment responses in resource-limited settings.

**Recommendation:** Given the exploratory nature of the findings, it is recommended that future studies investigate the role of inflammation/anti-inflammatory cytokine ratios, such as IL-17/IL-10, in larger and more diverse HIV-1 patient cohorts. Incorporating these biomarkers into clinical research may provide additional insights into immune regulation and treatment response, particularly in settings where viral load testing is limited. Validation of these cytokine ratios could support the development of cost-effective adjunct tools for HIV-1 monitoring and management in resource-constrained environments.

## Introduction

The global burden of HIV-1 remains high despite advances in combination antiretroviral therapy (cART). According to UNAIDS 2023, 39.0 million people lived with HIV-1 in 2022, with Eastern and Southeast Africa (ESA) accounting for 20.8 million cases(1). Kenya reported 1.4 million cases, with 18,000 AIDS-related deaths in the same year. Since the introduction of combination antiretroviral therapy (cART), Kenya has had 94% HIV-1 treatment coverage, with 1.2 million patients achieving viral suppression. Viral load suppression is critical in HIV-1 treatment, as sustained undetectable levels correlate with improved clinical outcomes and a reduced transmission risk (2). Despite this milestone, persistent low-level viremia (pLLV), defined as viral loads between 50-500 copies/ml, presents a challenge in HIV-1 patients (3). Persistent low-level viremia has been linked to virologic rebound (>1,000 copies/ml) and HIV-1 progression (4,5). Given limited viral load monitoring in Africa, understanding cytokine biomarker ratios in HIV-1 may offer alternative predictors of virologic rebound.

Interleukin-17 (IL-17) is a pro-inflammatory cytokine essential in host defense and immune regulation (6). In HIV-1 infection, IL-17 enhances mucosal immunity and cytotoxic responses via NK and CD8^+^ T cells (7), but its dysregulation contributes to mucosal barrier damage, immune activation, and chronic inflammation (8,9). IL-17 may also promote HIV-1 persistence by activating anti-apoptotic pathways in infected cells (10,11).

Interleukin-10 (IL-10) on the other hand is an anti-inflammatory cytokine produced by diverse immune cells including macrophages, dendritic cells, T cells, and B cells, and plays a critical role in immune homeostasis by suppressing pro-inflammatory cytokines, leading to inhibition of antigen-presenting cell function, and promoting regulatory T cell (Treg) differentiation and suppressive activity (12,13). In chronic infections such as HIV-1, elevated IL-10 levels dampen antiviral immunity by impairing dendritic cell activation, increasing the activation threshold for CD8^+^ cytotoxic T lymphocytes (CTLs), and attenuating their cytotoxic function, which collectively promote viral persistence and immune exhaustion (14,15). Through sustained immunosuppression, IL-10 disrupts effective CTL responses and facilitates HIV-1 immune evasion, contributing to progressive T cell dysfunction, chronic immune activation, and eventual progression to AIDS.

The IL-17/IL-10 ratio reflects the balance between pro- and anti-inflammatory responses, where a skew toward either direction may affect viral control, immune recovery, and long-term outcomes in HIV-1 infection. Despite its potential as a biomarker, its role in treatment monitoring remains underexplored in sub-Saharan Africa. Regional studies show persistent immune activation despite viral suppression, such as Kityo et al. (2017) in Uganda(16) and Musa et al., in Kenya(17), while Mlisana et al. (2012) reported dysregulated cytokine responses in early infection in South Africa (18) . Evaluating this ratio in HIV-1 patients at (Hospital), (Country), may offer valuable insights into immune dynamics in pLLV and inform cost-effective immune monitoring strategies in resource-limited settings.

This study assessed the viral load, as well as the IL17/IL10 ratio in virologically suppressed and pLLV patients on first-line cART to elucidate immune response differences and their role in HIV-1 rebound.

## Methodology

### Study population

The study population consisted of HIV-1 patients on first-line cART attending AMPATH Clinic, MTRH, Kenya, from January 2021 to December 2022. Test patients consisted of HIV-1 patients on first-line cART with pLLV with at least two consecutive episodes of VLs between 50-500 copies/ml in the past two years of the study. The comparison group consisted of HIV-1 patients on first-line cART, HIV-1 suppressed (LDL; VL<50 copies/ml).

### Participants Recruitment

A retrospective review of patient records from HIV-1 adult clinics identified individuals with post-suppression persistent low-level viremia (pLLV) and those with sustained viral suppression. Purposive sampling was used to deliberately select patients meeting the inclusion criteria-HIV-1– infected adults on first-line combination antiretroviral therapy (TDF + 3TC + DTG) with complete clinical and laboratory data. This was supplemented by consecutive sampling, where all eligible and available participants within the review period were included until the target sample size was attained. Eligible participants were contacted, informed about the study, and enrolled upon providing written consent. Matching of test and comparison groups by age and gender was conducted to ensure comparability and reduce potential confounding.

## Data Collection

### Sample Size Determination

The sample size was determined using the formula by Pagano and Gauvreau (2005):

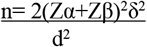

where *n* is the required sample size per group,

*Zα* is the desired level of statistical significance (1.96 for a 95% confidence interval),

*Zβ* represents the desired power (0.84),

*δ* is the standard deviation of the outcome variable of interest, and

*d* is the expected difference of the means between cases and controls.

The standard deviation (*δ*) was obtained from a previous related study (Osuji et al., 2018a), where transforming growth factor beta (TGF-β) had the highest mean of 61.24 ± 22.08. The expected mean difference between cases and controls was *d* = 13.8. Substituting these values into the formula gave:

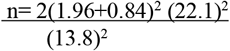

n = 41 per group

41 Cases and 41 for control

A total of 4,000 patient records were retrospectively reviewed to identify individuals meeting the inclusion criteria. After screening for eligibility, clinical completeness, and matching for age and gender, 41 patients with persistent low-level viremia (test group) and 41 with sustained viral suppression (control group) were enrolled upon providing informed consent.

For transparency, the standardized effect size used in the sample size calculation was computed as Cohen’s *d* = mean difference / standard deviation = 13.8 / 22.1 ≈ 0.62, corresponding to a moderate-to-large effect size. This assumed effect size, together with α = 0.05 and power = 0.80, informed the final sample size of 41 participants per group.

We recruited 82 HIV-1 patients on first-line cART, comprising 41 pLLV patients with 50–500 copies/ml and 41 age- and gender 1:1 matched, virally suppressed controls with <50 copies/ml. A qualified phlebotomist collected 4 mL blood samples for VL determination and cytokine analysis in EDTA tubes. The plasma was separated within 2 hours of collection and stored at -80°C until analysis day.

## Laboratory Procedures

### 1. ELISA for Cytokines

Serum concentrations of IL-17, IFN-γ, IL-10, and TGF-β were quantified using Genway’s Enzyme-Linked Immunosorbent Assay (ELISA) kits (Zeptometrix, Buffalo, NY, USA), following the manufacturer’s instructions with minor modifications to ensure procedural uniformity and accuracy.

Prior to analysis, the avidin–biotin enzyme complex (ABC) working solution and tetramethylbenzidine (TMB) substrate were equilibrated at 37°C for 30 minutes. Subsequently, 100 μL of standards and serum samples were dispensed into each well and incubated at 37°C for 90 minutes to allow antigen binding. Following this, biotinylated detection antibodies were added, and the plates were further incubated at 37°C for 60 minutes. Plates were washed three times with 0.01 M Tris-buffered saline (TBS) to remove unbound materials before adding the ABC working solution, which was incubated for another 30 minutes at 37°C.

After five additional washes with 0.01 M TBS, TMB substrate was added to each well, and color development was allowed to proceed until the reaction was stopped according to kit instructions. The optical density (OD) was then measured at 450 nm using a microplate reader within 30 minutes of adding the stop solution. Cytokine concentrations (pg/mL) were determined from a standard calibration curve generated using known concentrations of cytokine standards, and verified using Beer–Lambert’s law to confirm linearity between OD and concentration.

To ensure accuracy, precision, and reproducibility, all samples were analyzed in duplicate. Running samples in duplicates enabled identification and correction of any discrepancies arising from pipetting or technical variations. This duplication provided a built-in quality control measure that enhanced the reliability and validity of the cytokine quantification results.

### 1. PCR For Viral Load

Viral load was analyzed by polymerase chain reaction-Abbott Real-Time PCR HIV-1 RNA assay, version 4.0. A master mix with primers, nucleotides, buffer, and DNA polymerase was prepared. Template RNA was added, and thermal cycling was performed. PCR products were analyzed for target bands. Results were reported with amplification details.

The IL-17/IL-10 ratio was calculated to evaluate immune balance.

### Data Management and statistical analysis

Filled data forms were reviewed; incomplete ones were verified and refilled.Data were entered into Microsoft Excel® 2019 and then transferred to STATA version 15 for analysis. Normality was assessed with the Shapiro-Wilk test. The central tendency Median (IQR) measure was used to summarize cytokine levels and VL data. Mann-Whitney U-test was used to compare medians between the pLLV and HIV-1 suppressed patients. P<0.05 was considered statistically significant.

### Ethical Consideration

The study was approved by the Institutional Research and Ethics Committee of (Institution)-IREC approval-Ref. No. 0003744. For confidentiality, only the Clinical Officer identified participants from the HIV-1 adult clinic records. Informed consent was written by all patients who agreed to participate. Numeric codes were used to delink personal identifiers. Computer data was secured with password, while hard copies were secured in lockable cabinets, only accessible to the investigators.

## Results

Of 82 blood samples collected, 41 were form HIV-1 patients on first-line cART with persistent low-level viremia (pLLV) and 41 virologically suppressed HIV-1 patients, age and gender 1:1 matched. Participants were aged 18 to 57 years, 37 % (30) were between 38-47 years.

### Viral load in HIV-1 patients on first-line cART attending AMPATH clinic at MTRH

In HIV-1 patients on first-line cART with pLLV, the median (IQR) viral load was 407.5 (314.5– 445.0) copies/ml, while virologically suppressed patients had VL less than 50 copies/ml (not detectable) (Figure 1.)

**Figure 1:**
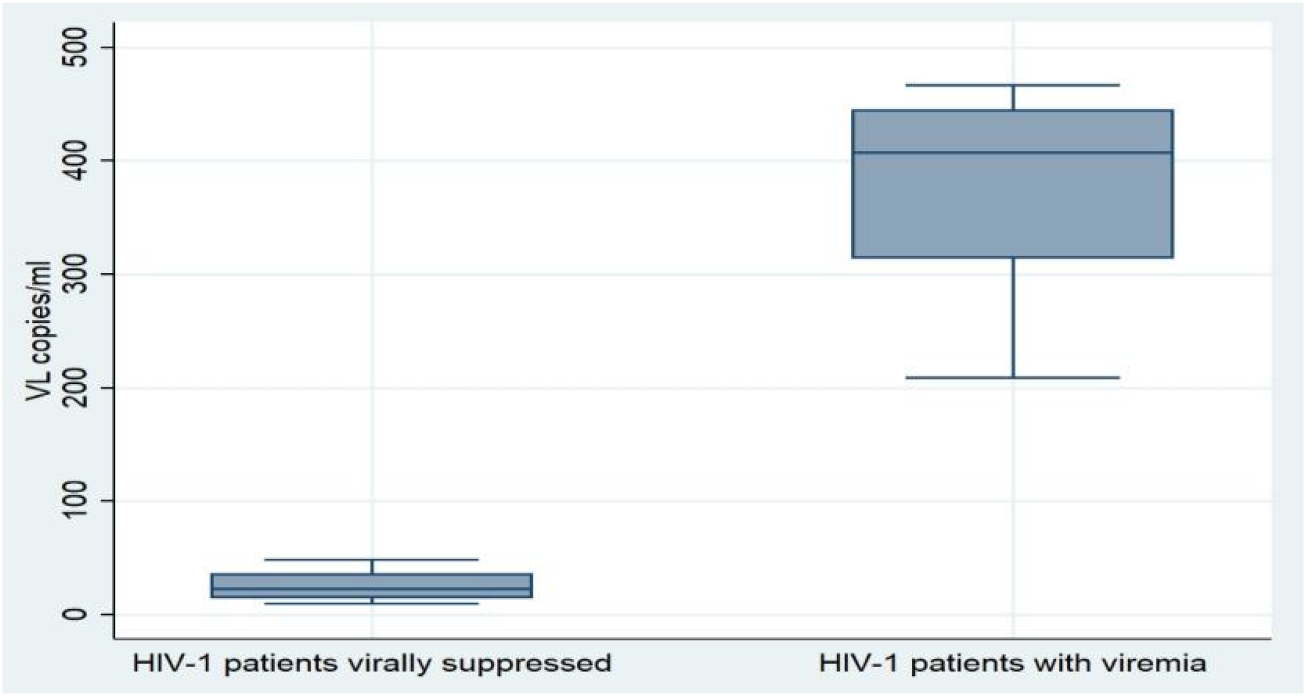
Median Viral load in HIV-1 patients on cART on first-line cART with persistent low-level viremia and patients on first-line cART, HIV-1 suppressed.

*The boxplot demonstrates a significant difference in viral load distributions between HIV-1-infected individuals on first-line combination antiretroviral therapy (cART) virally suppressed and those with persistent low-level viremia. Patients with viral suppression had VLs below the lower limit of quantification (<50 copies/ml), confirming effective treatment response. In contrast, viremic patients had a median VL of 407*.*5 copies/ml (IQR: 314*.*5–445*.*0), indicating ongoing viral replication despite cART*.

### Inflammatory/anti-inflammatory (IL-17/IL-10) ratios in HIV-1 patients with persistent low-level viremia and patients on first-line cART, HIV-1 suppressed on first-line cART

HIV-1-suppressed patients had a slightly higher mean IL-17/IL-10 ratio of 0.556 vs 0.552 in pLLV, P=0.433, and the ratio was positively correlated (rho= 0.453, p=0.003) to viral suppression in pLLV. Figure 2.

**Figure 2:**
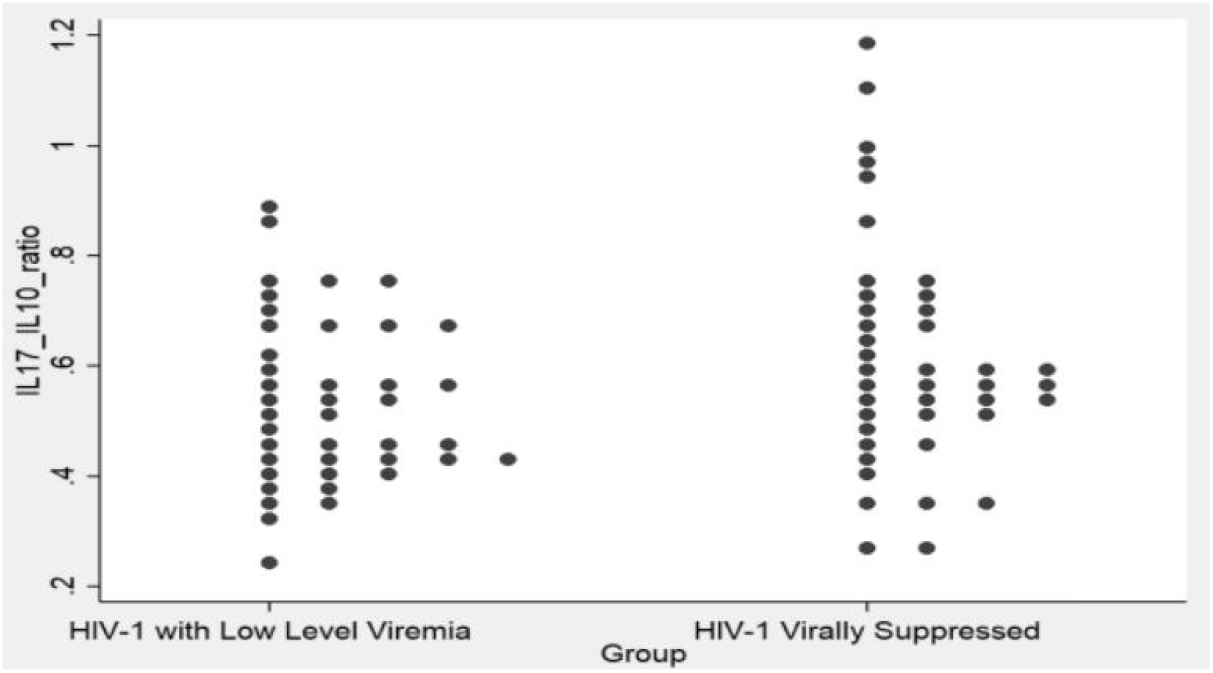
IL-17/IL-10 ratios in HIV-1 patients with persistent low-level viremia and HIV-1 patients on first-line cART who are virally suppressed on first-line cART.

*The dot plot illustrates the distribution of IL-17/IL-10 ratios in HIV-1 patients receiving first-line cART. The mean IL-17/IL-10 ratio was higher in virally suppressed patients (0*.*556) compared to those with persistent low-level viremia (pLLV) (0*.*552), although this difference was not statistically significant (P = 0*.*433). Within the pLLV group, the IL-17/IL-10 ratio demonstrated a moderate positive correlation with the degree of viral suppression (Spearman’s rho = 0*.*453, P = 0*.*003), suggesting a potential immunoregulatory role of this cytokine balance in controlling residual viremia*.

### Logistic regression of IL-17/IL-10 ratios on viremia

On simple logistic regression of IL-17/IL-10 ratios against viremia status as the dependent variable, we found that for each unit increase in the IL-17/IL-10 ratio, the odds of a patient being virally suppressed nearly double (OR 95% CI: 2.2; 0.3-17.9). However, this association was not statistically significant (p = 0.453), Table 1.

**Table 1.** Relationship between viremia and the IL-17/IL-10 ratio.

| Variable | Regression coefficient ($\beta$) | OR (95%CI) | P-value |
| --- | --- | --- | --- |
| IL-17/IL-10 ratio | 0.798 | 2.221(0.276-17.856) | 0.453 |
| Intercept | -0.488 | 0.614 |  |

## Discussion

### Viral load in HIV-1 patients on first-line cART attending AMPATH Clinic, MTRH

This study found some HIV-1 patients on first-line cART to have persistent low-level viremia with a median viral load of 407.5 copies/ml. Similarly, a study by Esber et al. found varying levels of viral load in HIV-1 patients receiving cART. Persistent low-level viremia was detected in 20% of patients, while 2% of the patients in the study had PLLV ranging from 200 to 999 copies/ml (20). This current study’s findings reveal varying levels of viral load, 50-500 copies/ml, among HIV-1 patients on first-line cART with persistent low-level viremia. This finding supports earlier findings that even with seemingly effective cART treatment, viral loads can fluctuate above the detection threshold of >50 copies/ml among individuals living with HIV-1 on cART. While the primary treatment goal for patients with HIV-1 on cART is to achieve and maintain an undetectable viral load (21), it is recognized that persistent low-level viremia can still occur in some patients (3,22). The viral load observed in the current study may be indicative of ongoing, albeit low-level, viral replication despite treatment. This persistence of the virus reflects incomplete viral suppression or the existence of reservoirs in the body where the virus remains and is less affected by the current treatment regimen. Additionally, latent HIV-1 in immune cells, such as resting CD4+ T cells, may evade detection and treatment, reactivating to cause low-level viral replication. The observed fluctuations could also mean that certain factors, such as drug resistance or suboptimal drug penetration in specific tissues, might contribute to viral persistence. In contrast to the current study, Young and colleagues (23) reported a consistent decrease in the proportion of patients experiencing an unsuccessful virologic outcome throughout cART. This suggests that the success of cART may have contributed to the favorable virologic outcomes observed in their study, accounting for the contrasting results. The success of cART was due to its ability to consistently suppress HIV replication, reduce viral load, and restore immune function, leading to improved health outcomes. Immune activation and co-infections, such as hepatitis C, can further facilitate viral persistence by facilitating inflammation. Also, if viral loads exceed 50 copies/ml, it may indicate treatment inefficacy, possibly due to resistance or adherence issues, increasing the risk of virologic failure (20,24). The varying viral loads observed in the current study suggest ongoing replication, which could lead to the accumulation of drug-resistant mutations and eventual treatment failure.

### IL-17/ IL-10 ratios in HIV-1 patients on first-line cART with pLLV and virally suppressed patients

This study found slightly higher mean IL-17/IL-10 ratios in HIV-1-suppressed patients on first-line cART (0.556) compared to those with persistent low-level viremia (0.552); though not statistically significant (Z = 0.784, P = 0.433). The IL-17/IL-10 ratio reflects the balance between pro-inflammatory (IL-17) and anti-inflammatory (IL-10). A higher ratio indicates more inflammation relative to immune regulation due to anti-inflammatory cytokines. In HIV-1-suppressed patients on first-line cART, the ratio is slightly higher (0.556) compared to those with persistent low-level viremia, suggesting ongoing immune dysregulation despite viral suppression.

Persistent low-level viremia may involve a more balanced immune response with higher IL-10 to control inflammation. The higher mean IL-17/IL-10 ratio observed in suppressed patients suggests an altered immune response, potentially indicating a shift towards a more pro-inflammatory state. This could reflect an adaptive immune reaction to HIV-1 control or an indicator of immune system rebalancing following effective viral suppression (8). Similar findings have been reported by Falivene et al. (25), who found an imbalanced ratio in HIV-1 patients on cART, suggesting improved infection control. The elevated IL-17/IL-10 ratio in HIV-1 patients on cART suggests improved viral control by pro-inflammatory cytokines such as IL-17, which play a role in recruiting other immune cells to control the viral infection. In contrast, IL-10, an anti-inflammatory cytokine, helps control excessive immune responses and disease progression. A higher mean IL-17/IL-10 ratio in these patients thus indicates, despite viral suppression, that the immune system remains more responsive, boosting defense against residual HIV-1. This enhanced immune response helps control HIV-1 replication and limit disease progression, reflecting a crucial balance between inflammation and immune regulation for effective long-term management. In contrast, Yanshuang and colleagues (26) reported a lower IL-17/IL-10 ratio in HIV-1 patients co-infected with tuberculosis, with HIV-1 replication and disease progression. Co-infections like tuberculosis in HIV-1 patients likely lead to a lower IL-17/IL-10 ratio due to immune dysregulation, where HIV-1 weakens immune responses and TB induces chronic inflammation, resulting in reduced IL-17 production and an increased compensatory IL-10 response to control inflammation. The IL-17/IL-10 balance plays a crucial role in HIV-1 infection, influencing viral control, immune activation, inflammation, tissue damage, and viral persistence. Its significance extends beyond pathogenesis, as it may enhance treatment response and serve as a valuable prognostic biomarker for disease progression and therapeutic outcomes (26).

## Conclusion

Viral suppression in HIV-1 patients on cART is related to inflammatory/anti-inflammatory IL-17/IL-10 ratios in HIV-1 Patients on First-Line Antiretroviral Therapy at Moi Teaching and referral Hospiral, Kenya.

### Recommendation

The study found no statistically significant difference in IL-17/IL-10 ratios between pLLV and virally suppressed HIV-1 patients on cART. While these findings suggest a possible involvement of IL-17/IL-10 immune balance in viral control, they remain exploratory. Further research with larger cohorts is needed to clarify these relationships and to determine whether cytokine biomarkers could complement existing tools for monitoring treatment responses in resource-limited settings.

### Study Limitations

Other factors can influence cytokine profiles and their roles in the immune system. Comorbidities, immune disorders, and other confounders could affect the levels of cytokines found in a patient. It was impossible to verify the self-reported comorbidities, which could negatively impact the results found. The time duration since the initiation of cART was not factored in, and this could have a variation in the findings of the research.

### Explanation for Not Adjusting for Potential Confounders

Potential confounders such as duration on cART and treatment adherence were not adjusted for in the analysis due to incomplete and inconsistent documentation in patient medical records. As this was a retrospective study, key variables related to treatment timelines and adherence monitoring were either missing or not uniformly recorded across participants, making statistical adjustment unreliable. Including such incomplete data could have introduced bias or compromised the validity of the results. Consequently, these variables were acknowledged as unmeasured confounders and discussed under the *Study Limitations* section.

## Data Availability

All data produced in the present study are available upon reasonable request to the authors

## Acknowledgment

The ALMIGHTY GOD for the grace and favor. All staff in the Immunology Section, Department of Pathology, Moi University.

## Author Approvals

All authors have seen and approved the manuscript. This manuscript has not been accepted or published elsewhere.

## References

1. The Path That Ends AIDS: 2023 UNAIDS Global AIDS Update [Internet]. [cited 2023 Sep 28]. Available from: https://repository.gheli.harvard.edu/repository/collection/resource-pack-hiv/resource/12120/

2. Montaner JSG, Lima VD, Harrigan PR, Lourenço L, Yip B, Nosyk B, et al. Expansion of HAART Coverage Is Associated with Sustained Decreases in HIV/AIDS Morbidity, Mortality and HIV Transmission: The “HIV Treatment as Prevention” Experience in a Canadian Setting. PLOS ONE [Internet]. 2014 Feb 12 [cited 2023 Nov 7];9(2):e87872. Available from: https://journals.plos.org/plosone/article?id=10.1371/journal.pone.0087872

3. Elvstam O, Medstrand P, Yilmaz A, Isberg PE, Gisslén M, Björkman P. Virological failure and all-cause mortality in HIV-positive adults with low-level viremia during antiretroviral treatment. PLOS ONE [Internet]. 2017 Jul 6 [cited 2023 Apr 24];12(7):e0180761. Available from: https://journals.plos.org/plosone/article?id=10.1371/journal.pone.0180761

4. Arpadi SM, Shiau S, Gusmao EPD, Violari A. Routine viral load monitoring in HIV-infected infants and children in low- and middle-income countries: challenges and opportunities. Journal of the International AIDS Society [Internet]. 2017 Nov 24 [cited 2023 Nov 7];20:e25001. Available from: https://onlinelibrary.wiley.com/doi/10.1002/jia2.25001

5. Quiros-Roldan E, Raffetti E, Castelli F, Focà E, Castelnuovo F, Di Pietro M, et al. Low-level viraemia, measured as viraemia copy-years, as a prognostic factor for medium–long-term all-cause mortality: a MASTER cohort study. J Antimicrob Chemother [Internet]. 2016 Dec 1 [cited 2023 Oct 11];71(12):3519–27. Available from: https://dx.doi.org/10.1093/jac/dkw307

6. Jin W, Dong C. IL-17 cytokines in immunity and inflammation. Emerging Microbes & Infections [Internet]. 2013 Jan 1 [cited 2023 Nov 1]; Available from: https://www.tandfonline.com/doi/abs/10.1038/emi.2013.58

7. Mills KHG. IL-17 and IL-17-producing cells in protection versus pathology. Nat Rev Immunol [Internet]. 2023 Jan [cited 2023 Sep 23];23(1):38–54. Available from: https://www.nature.com/articles/s41577-022-00746-9

8. McGeachy MJ, Cua DJ, Gaffen SL. The IL-17 Family of Cytokines in Health and Disease. Immunity [Internet]. 2019 Apr [cited 2023 Aug 11];50(4):892–906. Available from: https://linkinghub.elsevier.com/retrieve/pii/S1074761319301384

9. Abusleme L, Moutsopoulos NM. IL-17: overview and role in oral immunity and microbiome. Oral Diseases [Internet]. 2017 Oct 1 [cited 2024 Feb 19];23(7):854–65. Available from: https://onlinelibrary.wiley.com/doi/10.1111/odi.12598

10. Fang Y, Peng K. Regulation of innate immune responses by cell death-associated caspases during virus infection. The FEBS Journal [Internet]. 2022 Jul 1 [cited 2024 Feb 19];289(14):4098–111. Available from: https://febs.onlinelibrary.wiley.com/doi/10.1111/febs.16051

11. Kuwabara T, Ishikawa F, Kondo M, Kakiuchi T. The Role of IL-17 and Related Cytokines in Inflammatory Autoimmune Diseases. Mediators of Inflammation [Internet]. 2017 Feb 20 [cited 2024 Feb 19];2017. Available from: https://www.hindawi.com/journals/mi/2017/3908061/

12. Rutz S, Ouyang W. Regulation of Interleukin-10 Expression. In: Regulation of Cytokine Gene Expression in Immunity and Diseases [Internet]. Springer, Dordrecht; 2016 [cited 2024 Feb 20]. p. 89–116. Available from: https://link.springer.com/chapter/10.1007/978-94-024-0921-5_5

13. Saraiva M, Vieira P, O’Garra A. Biology and therapeutic potential of interleukin-10. J Exp Med [Internet]. 2020 Jan 6 [cited 2023 Apr 25];217(1). Available from: https://rupress.org/jem/article/217/1/e20190418/132577/Biology-and-therapeutic-potential-of-interleukin

14. Lobo-Silva D, Carriche GM, Castro AG, Roque S, Saraiva M. Balancing the immune response in the brain: IL-10 and its regulation. J Neuroinflammation [Internet]. 2016 Dec [cited 2024 Feb 20];13(1):1–10. Available from: https://jneuroinflammation.biomedcentral.com/articles/10.1186/s12974-016-0763-8

15. Rojas JM, Avia M, Martín V, Sevilla N. IL-10: A Multifunctional Cytokine in Viral Infections. Journal of Immunology Research [Internet]. 2017 Feb 20 [cited 2023 Apr 25];2017. Available from: https://www.hindawi.com/journals/jir/2017/6104054/

16. Dirajlal-Fargo S, Strah M, Ailstock K, Sattar A, Karungi C, Nazzinda R, et al. Persistent immune activation and altered gut integrity over time in a longitudinal study of Ugandan youth with perinatally acquired HIV. Front Immunol [Internet]. 2023 Mar 28 [cited 2025 May 1];14:1165964. Available from: https://www.frontiersin.org https://www.frontiersin.org/journals/immunology/articles/10.3389/fim mu.2023.1165964/full

17. Musa F, Shaviya N, Mambo F, Abonyo C, Barasa E, Wafula P, et al. Cytokine profiles in highly active antiretroviral treatment non-adherent, adherent and naive HIV-1 infected patients in Western Kenya. 1 [Internet]. 2021 Dec 14 [cited 2023 Apr 24];21(4):1584–92. Available from: https://www.ajol.info/index.php/ahs/article/view/218807

18. Roberts L, Passmore JAS, Williamson C, Little F, Bebell LM, Mlisana K, et al. Plasma cytokine levels during acute HIV-1 infection predict HIV disease progression. AIDS (London, England) [Internet]. 2010 Mar 3 [cited 2023 Apr 28];24(6):819. Available from: https://www.ncbi.nlm.nih.gov/pmc/articles/PMC3001189/

19. Pagano M, Gauvreau K. Principles of Biostatistics. 2nd edition. Pacific Grove, CA: Duxbury Press; 2000. 592 p.

20. Esber A, Polyak C, Kiweewa F, Maswai J, Owuoth J, Maganga L, et al. Persistent Low-level Viremia Predicts Subsequent Virologic Failure: Is It Time to Change the Third 90? Clin Infect Dis [Internet]. 2019 Aug 16 [cited 2023 Apr 24];69(5):805–12. Available from: https://academic.oup.com/cid/article/69/5/805/5193460

21. Hofstra LM, Mudrikova T, Stam AJ, Otto S, Tesselaar K, Nijhuis M, et al. Residual Viremia Is Preceding Viral Blips and Persistent Low-Level Viremia in Treated HIV-1 Patients. PLOS ONE [Internet]. 2014 Oct 29 [cited 2023 Oct 11];9(10):e110749. Available from: https://journals.plos.org/plosone/article?id=10.1371/journal.pone.0110749

22. Crespo-Bermejo C, Arellano ER de, Lara-Aguilar V, Valle-Millares D, Gómez-Lus ML, Madrid R, et al. Persistent low-Level viremia in persons living with HIV undertreatment: An unresolved status. Virulence [Internet]. 2021 Dec 7 [cited 2023 Apr 24]; Available from: https://www.tandfonline.com/doi/abs/10.1080/21505594.2021.2004743

23. Young PW, Musingila P, Kingwara L, Voetsch AC, Zielinski-Gutierrez E, Bulterys M, et al. HIV Incidence, Recent HIV Infection, and Associated Factors, Kenya, 2007–2018. AIDS Research and Human Retroviruses [Internet]. 2023 Feb 8 [cited 2024 Feb 20]; Available from: https://www.liebertpub.com/doi/10.1089/aid.2022.0054

24. Gaifer Z, Boulassel MR. Low-Level Viremia Predicts Virological Failure in HIV-Infected Omani Patients Receiving Antiretroviral Therapy. J Int Assoc Provid AIDS Care [Internet]. 2020 Jan 1 [cited 2023 Apr 24];19:232595822097981. Available from: http://journals.sagepub.com/doi/10.1177/2325958220979817

25. Falivene J, Ghiglione Y, Laufer N, Socías ME, Holgado MP, Ruiz MJ, et al. Th17 and Th17/Treg ratio at early HIV infection associate with protective HIV-specific CD8^+^ T-cell responses and disease progression. Sci Rep [Internet]. 2015 Jun 23 [cited 2024 Jun 27];5(1):11511. Available from: https://www.nature.com/articles/srep11511

26. Li Y, Sun W. Effects of Th17/Treg cell imbalance on HIV replication in patients with AIDS complicated with tuberculosis. Experimental and Therapeutic Medicine [Internet]. 2018 Mar 1 [cited 2023 Apr 30];15(3):2879–83. Available from: https://www.spandidos-publications.com/10.3892/etm.2018.5768

